# Level of IL-6 predicts respiratory failure in hospitalized symptomatic COVID-19 patients

**DOI:** 10.1101/2020.04.01.20047381

**Authors:** Tobias Herold, Vindi Jurinovic, Chiara Arnreich, Johannes C. Hellmuth, Michael von Bergwelt-Baildon, Matthias Klein, Tobias Weinberger

## Abstract

The pandemic Coronavirus-disease 19 (COVID-19) is characterized by a heterogeneous clinical course. While most patients experience only mild symptoms, a relevant proportion develop severe disease progression with increasing hypoxia up to acute respiratory distress syndrome. The substantial number of patients with severe disease have strained intensive care capacities to an unprecedented level. Owing to the highly variable course and lack of reliable predictors for deterioration, we aimed to identify variables that allow the prediction of patients with a high risk of respiratory failure and need of mechanical ventilation Patients with PCR proven symptomatic COVID-19 infection hospitalized at our institution from 29th February to 27th March 2020 (n=40) were analyzed for baseline clinical and laboratory findings. Patients requiring mechanical ventilation 13/40 (32.5%) did not differ in age, comorbidities, radiological findings, respiratory rate or qSofa score. However, elevated interleukin-6 (IL-6) was strongly associated with the need for mechanical ventilation (p=1.2.10^-5^). In addition, the maximal IL-6 level (cutoff 80 pg/ml) for each patient during disease predicted respiratory failure with high accuracy (p=1.7.10^-8^, AUC=0.98). The risk of respiratory failure for patients with IL-6 levels of >; 80 pg/ml was 22 times higher compared to patients with lower IL-6 levels. In the current situation with overwhelmed intensive care units and overcrowded emergency rooms, correct triage of patients in need of intensive care is crucial. Our study shows that IL-6 is an effective marker that might be able to predict upcoming respiratory failure with high accuracy and help physicians correctly allocate patients at an early stage.

## Introduction

The pandemic Coronavirus-disease 19 (COVID-19) is characterized clinically by a highly variable course. While most patients experience only mild symptoms, a relevant proportion develop severe disease progression with increasing hypoxia up to acute respiratory distress syndrome. About 5% of patients require intensive care treatment including mechanical ventilation. ^1-3^ This variability of COVID-19 and the shortage of health care resources in heavily affected regions make efficient allocation of resources towards patients at high risk for deterioration crucial.^4^ We aimed to identify variables that allow the prediction of patients with a high risk of respiratory failure and need of mechanical ventilation.

## Methods

All patients with PCR proven symptomatic COVID-19 infection hospitalized at our institution from 29^th^ February to 27^th^ March 2020 (n=40) were analyzed for baseline clinical and laboratory findings. Patient data were anonymized for analysis and the study was approved by the local ethics committee (No: 20-245). All variables with less than 50% of missing data were tested for the association with respiratory failure. Categorical variables were compared with the χ^2^-test, and numerical variables were tested with the Mann-Whitney-U-Test. The p-values were adjusted for multiple testing with the Bonferroni-Holm-Method. An adjusted p-value (q-value) of ≤0.05 was considered significant.

## Results

In total, 13/40 (32.5%) patients deteriorated during hospitalization and required mechanical ventilation. The time from hospital admission to intubation varied from less than one hour to 9 days (median 2 days). All patients who required intubation were of male sex, compared to 59% of males in the group that did not require intubation (p=0.020, q=0.057). Patients requiring mechanical ventilation did not differ in age, comorbidities, radiological findings, respiratory rate or qSofa score (table).

**Table.**

| Variable | Evaluable | Median (range) /<br>n (%) | Mechanical ventilation |  | p-value | q-value |
| --- | --- | --- | --- | --- | --- | --- |
|  |  |  | No (n = 27) | Yes (n = 13) |  |  |
| Baseline Characteristics |  |  |  |  |  |  |
| Age (years) | 40 | 57 (19 - 81) | 54 (19 - 80) | 64 (45 - 81) | 0.15 | 0.28 |
| Respiratory rate (/min) | 33 | 18 (14 - 40) | 18 (14 - 32) | 23 (15 - 40) | 0.075 | 0.15 |
| Pulse (/min) | 33 | 82 (54 - 130) | 77 (54 - 111) | 95 (80 - 130) | **0.0028** | **0.012** |
| Male gender | 40 | 29 (72) | 16 (59) | 13 (100) | **0.020** | 0.058 |
| Any comorbidities | 39 | 32 (82) | 20 (77) | 12 (92) | 0.46 | 0.70 |
| Hypertension | 36 | 19 (53) | 10 (40) | 9 (82) | 0.051 | 0.11 |
| Diabetes mellitus | 37 | 3 (8) | 1 (4) | 2 (17) | 0.50 | 0.72 |
| Coronary heart disease | 34 | 3 (9) | 3 (12) | 0 (0) | 0.61 | 0.80 |
| Chronic obstructive lung disease | 35 | 3 (9) | 2 (8) | 1 (9) | >0.99 | >0.99 |
| Computed Tomography |  |  |  |  |  |  |
| Consolidation | 35 | 20 (57) | 14 (61) | 6 (50) | 0.80 | 0.99 |
| Ground glass opacity | 35 | 31 (89) | 20 (87) | 11 (92) | >0.99 | >0.99 |
| Bilateral infiltration | 35 | 31 (89) | 21 (91) | 10 (83) | 0.89 | >0.99 |
| Scores |  |  |  |  |  |  |
| qSOFA score | 31 | 12 (39) | 7 (32) | 5 (56) | 0.41 | 0.66 |
| CURB-65 score ≥1 | 23 | 7 (30) | 5 (31) | 2 (29) | >0.99 | >0.99 |
| Laboratory parameters | Evaluable | Median (range) | Mechanical ventilation |  | p-value | q-value |
|  |  |  | No (n = 27) | Yes (n = 13) |  |  |
| Lymphocyte count (%) | 36 | 19 (4 - 45) | 21 (4 - 45) | 15 (6 - 26) | **0.050** | 0.11 |
| CRP (mg/dl) | 40 | 2.8 (0 – 31.5) | 1.7 (0 – 31.5) | 7.8 (1.6 – 17.1) | **0.0019** | **0.012** |
| Bilirubin (mg/dl) | 36 | 0.5 (0.2 – 1.9) | 0.5 (0.2 – 1.2) | 0.5 (0.4 – 1.9) | 0.93 | >0.99 |
| WBC (G/l) | 40 | 5.295 (2.12 - 308) | 4.75 (2.12 – 12.5) | 6.64 (4.99 - 308) | **0.0014** | **0.012** |
| LDH (U/l) | 38 | 292 (182 - 1078) | 281 (182 - 619) | 346 (252 - 1078) | **0.0026** | **0.012** |
| PCT (ng/ml) | 37 | 0 (0 - 5) | 0 (0 – 0.6) | 0.1 (0 - 5) | **0.011** | **0.041** |
| IL6 (pg/ml) | 37 | 27.1 (0 - 430) | 19.6 (0 – 76.5) | 121 (19.2 - 430) | **0.000012** | **0.00032** |
| Thrombocyte count (G/l) | 40 | 165 (88 - 440) | 186 (88 - 334) | 160 (1 - 440) | 0.59 | 0.80 |
| Troponin T (ng/ml) | 34 | 0 (0 – 0.032) | 0 (0 – 0.022) | 0 (0 – 0.032) | **0.018** | 0.058 |
| Creatinine (mg/dl) | 40 | 0.9 (0.4 – 2.1) | 0.9 (0.4 – 1.3) | 1.0 (0.9 – 2.1) | **0.00034** | **0.0044** |
| D-Dimer | 30 | 0.7 (0 – 2.9) | 0.6 (0 – 2.2) | 1.1 (0.6 – 2.9) | **0.028** | 0.074 |
| Ferritin (ng/ml) | 27 | 644 (64 - 2153) | 606 (64 - 1748) | 810 (431 - 2153) | 0.16 | 0.28 |

CRP = C-Reactive Protein; WBC= White blood cell count; LDH = Lactate Dehydrogenase; PCT = Procalcitonin; IL6 = Interleukin-6

Pulse, markers of inflammation, LDH and creatinine at admission were associated with respiratory failure (table).

Elevated interleukin-6 (IL-6) was very strongly associated with the need for mechanical ventilation (figure 1A, p=1.2·10^−5^, q=0.00032).

**Figure 1:**
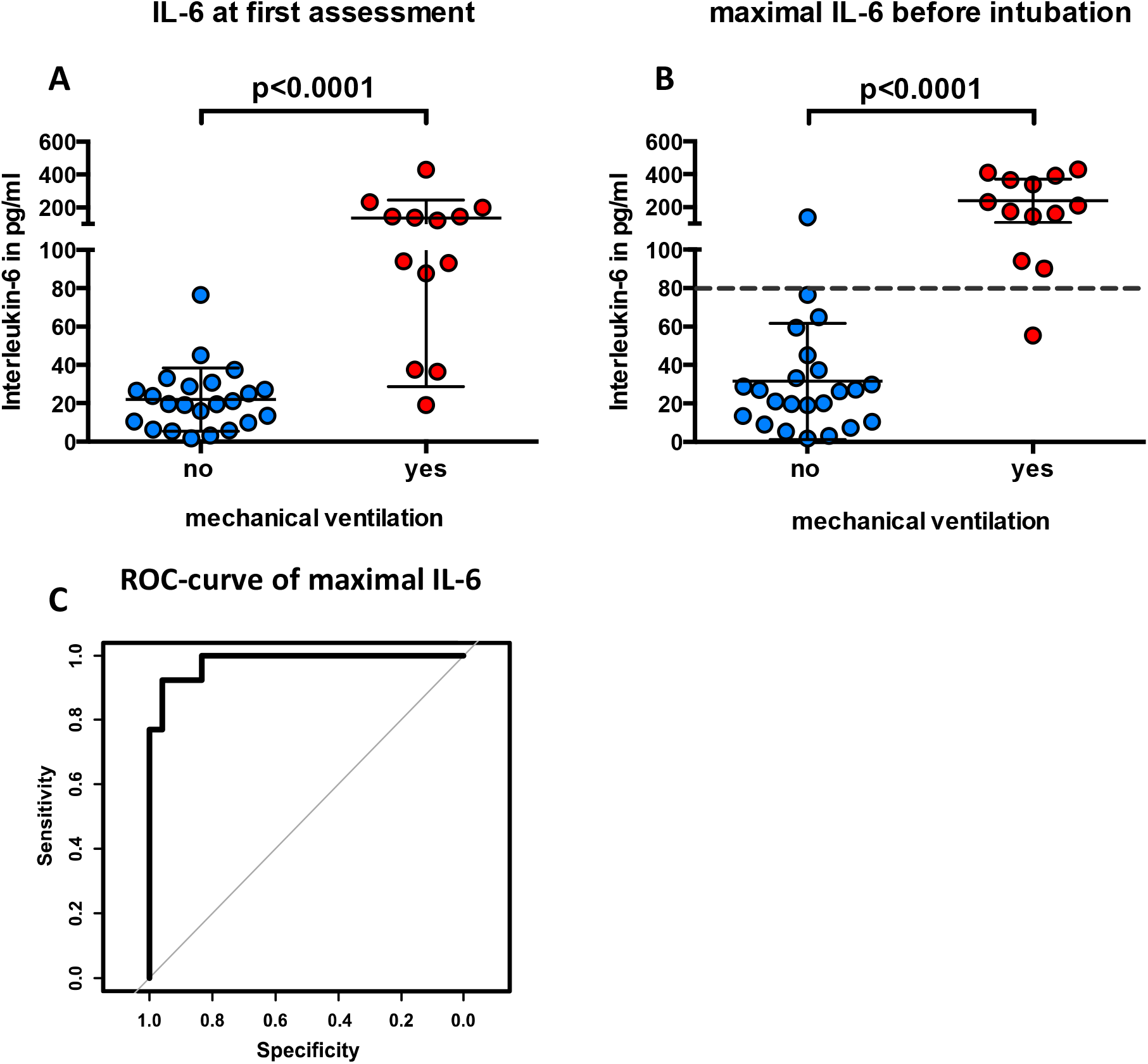
Box plot showing IL-6 levels at first assessment p=1.2·10^−5^ (A) and maximal IL-6 levels before mechanical ventilation p=1.7·10^−8^ (B). Receiver operating characteristic (ROC) curve of maximal IL-6 levels before mechanical ventilation, AUC=0.98 (C).

In addition to IL-6 values at first assessment, follow-up data on IL-6 were also available (median number of values per patient: 6, range 1-10). These data were used to assess the maximal IL-6 level for each patient during disease (for patients requiring ventilation, only values before intubation were used). These values predicted respiratory failure with high accuracy (figure 1B/C, p=1.7·10^−8^, AUC=0.98). The statistically optimal cutoff for IL-6 was 80pg/ml, identifying all but one patient with respiratory failure correctly, and misclassifying only one patient that still does not require intubation. The risk of respiratory failure for patients with IL-6 levels of ≥80pg/ml was 92% and thus 22 times higher compared to patients with lower IL-6 levels. After reaching an IL-6 value of 80pg/ml, the median time to mechanical ventilation was 1.5 days (range 0–4 days).

## Discussion

Even though IL-6 levels were significantly elevated in patients requiring ventilation, they are relatively low compared to levels observed in patients with septic shock. ^5^ Our data suggests that even moderately elevated IL-6 levels above 80pg/ml are sufficient to identify COVID-19 patients with a high risk of respiratory failure. Further studies and larger sample sizes will be needed to validate our findings and possibly determine a more accurate cutoff. To date, it is unclear whether IL-6 merely represents a biomarker or a central pathogenetic element of severe COVID-19 that should be used as a parameter for therapeutic intervention.

In the current situation with overwhelmed intensive care units and overcrowded emergency rooms, correct triage of patients in need of intensive care is crucial. Our study shows that IL-6 is an effective marker that might be able to predict upcoming respiratory failure with high accuracy and help physicians correctly allocate patients at an early stage.

## Data Availability

We note that patient data can be made available from the authors upon reasonable request and with permission of the local ethics committee.

## Acknowledgement

We thank Oliver T. Keppler, the Task Force Corona at the University Hospital, LMU Munich and all CORKUM investigators as well as all health care workers for their outstanding service.

